# Ngari virus (Orthobunyavirus, Peribunyaviridae) in ixodid ticks collected from cattle in Guinea

**DOI:** 10.1101/2020.11.10.20228924

**Authors:** MT Makenov, AH Toure, RB Bayandin, AV Gladysheva, AV Shipovalov, S Boumbaly, N Sacko, MG Korneev, SA Yakovlev, OB Zhurenkova, YaE Grigoreva, MV Fyodorova, EV Radyuk, ES Morozkin, MY Boiro, K Khafizov, A Matsvay, LS Karan

## Abstract

Ngari virus is a mosquito-borne virus belonging to the genus Orthobunyavirus (Peribunyaviridae family). This virus is pathogenic to humans and causes severe illness. Ngari virus is present in several African countries, including Madagascar. Here, we report the detection of Ngari virus in ixodid ticks collected from cows in Guinea.

A tick survey was conducted in March-November of 2018 in six regions of Guinea. The sample comprised 710 pools, with a total of 2067 ticks belonging to five species collected from 197 cows. At the initial stage, we screened a subsample of tick pools of vector-borne viruses with a multiplex genus-specific primer panel. In the second stage of the study, we narrowed the search and screened all the samples by qPCR for the detection of Ngari virus. All positive samples were sequenced with primers flanking Ngari virus-specific fragments on the S and M segments.

We found Ngari virus in 12 pools that were formed from engorged ticks collected from livestock in three villages of the Kindia and Kankan regions. Sequencing of the S and M segments confirmed that the detected viruses belong to Ngari virus, and the viruses were most similar to the strain Adrar, which was isolated in Mauritania. We detected viral RNA in ticks of the following species: *Amblyomma variegatum, Rhipicephalus geigyi*, and *Rh. (Boophilus) spp*. There is no evidence that ixodid ticks are competent vectors of the Ngari virus. Most likely, the ticks obtained the virus through blood from an infected host.

## Introduction

Ngari virus is a mosquito-borne virus of the *Orthobunyavirus* genus (Peribunyaviridae family) (Abudurexiti et al., 2019). Currently, Ngari virus is classified as *Bunyamwera orthobunyavirus* (Abudurexiti et al., 2019). The viruses of the Peribunyaviridae family have a segmented genome that consists of three segments: small, medium and large segments (Elliott, 2014). Ngari virus is a natural reassortant: the L and S segments are from *Bunyamwera orthobunyavirus*, and the M segment is from *Batai orthobunyavirus* (Bowen et al., 2001; Gerrard et al., 2004). Ngari virus is pathogenic to humans and causes severe and fatal hemorrhagic fever (Gerrard et al., 2004). A range of mosquito species of the *Aedes, Anopheles*, and *Culex* genera are able to transmit this virus (Dutuze et al., 2018). Besides mosquitoes, the Ngari virus has also been found in cows and small ruminants (Dutuze et al., 2020; Eiden et al., 2014). To date, Ngari virus has been detected in countries of Sub-Saharan Africa, including Senegal, Mauritania, Burkina Faso, the Central African Republic, the Democratic Republic of the Congo, Sudan, Kenya, Somalia, South Africa, and Madagascar (Dutuze et al., 2018). Febrile illnesses in humans caused by Ngari virus have been confirmed in Sudan, Somalia, and Kenya (Bowen et al., 2001; Briese et al., 2006).

In Guinea, the Ngari virus has not been reported previously. However, there are a few reports of the presence of other Orthobunyaviruses in Guinea (Butenko, 1996; Jentes et al., 2010). As part of a joint long-term project of Guinea and the USSR, a large virological work was carried out in Guinea from 1978-1989, during which 127 strains of arboviruses of 20 species were obtained (Butenko, 1996). Most of them were found in ixodid ticks. This list includes four strains of *Bunyamwera orthobunyavirus* and four strains of *M’Poko orthobunyavirus* identified using serological methods (Butenko, 1996). Later, IgM antibodies for Tahyna virus were detected in the serum of five patients with acute infection in Guinea (Jentes et al 2010). These findings were confirmed by virus-specific neutralizing antibody 90% titer endpoint plaque-reduction neutralization (PRNT) assays. Furthermore, the authors described a case that was positive on ELISA IgM for Tahyna virus but was PRNT negative, presumably indicating infection with a different bunyavirus (Jentes et al 2010). Thus, previous works showed the presence of Orthobunyaviruses in Guinea. However, the virus species were identified using virological and/or serological methods only, and the obtained isolates were not sequenced. Taking into account these findings, we aimed to screen ticks in Guinea for the presence of different vector-borne pathogens. Here, we report the detection of Ngari virus in ixodid ticks collected from cows in Guinea.

## Methods

### Study design

A tick survey was conducted in March-November of 2018 in six regions of Guinea: Kindia, Boke, Mamou, Faranah, Kankan, and N’Zerekore (Fig. 1). The sample encompassed 710 pools, with a total of 2067 ticks belonging to five species collected from 197 cows (Table 1).

**Table 1.**
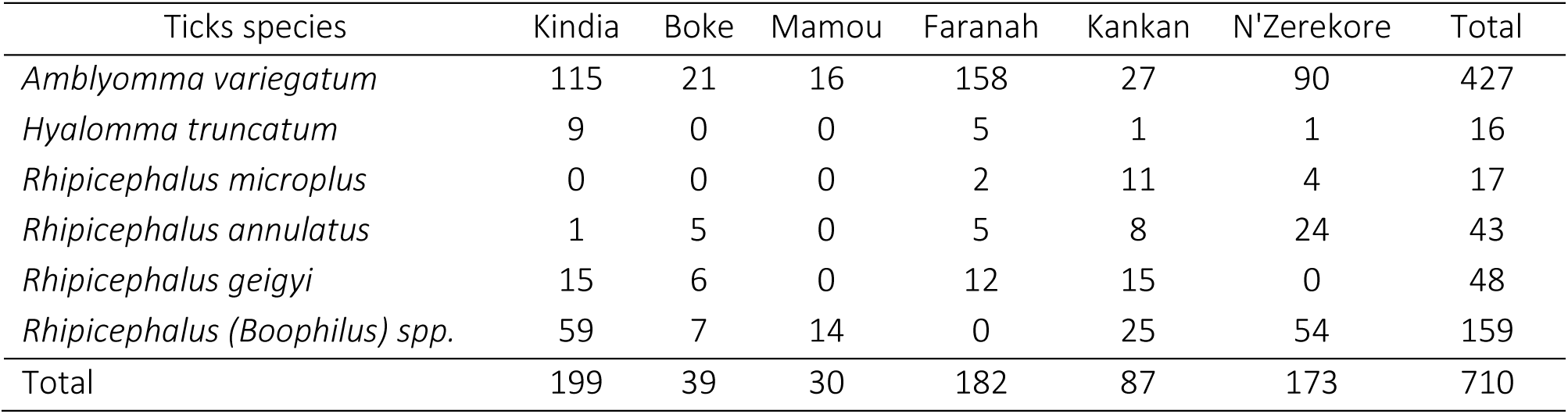
Total number of tick pools screened for the presence of Ngari virus

**Fig.1.**
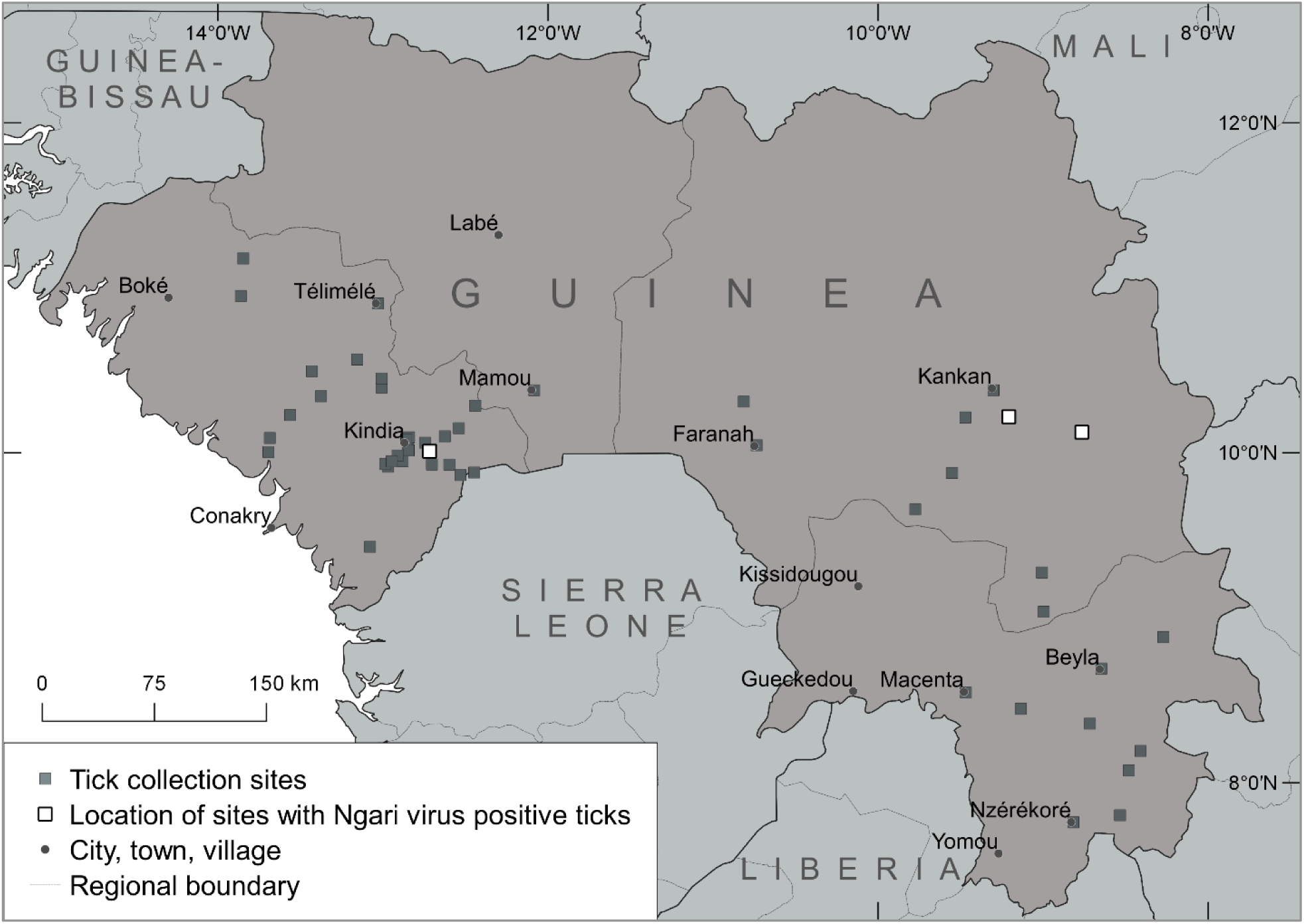
Map of collection sites with depicted locations of Ngari-positive tick detection

At the initial stage, we screened a subsample of tick pools for vector-borne viruses with a multiplex genus-specific primer panel (see below for a more detailed description). The subsample included specimens that had already been identified and homogenized in June 2018. The subsample encompassed 167 pools, with a total of 254 ticks collected from 65 animals in April-May 2018 in five out of six studied regions of Guinea (Table 2). During subsample testing, we found the Ngari virus in four pools, and subsequently, we focused our subsequent research on Ngari virus screening.

**Table 2.** Number of tick pools of vector-borne viruses screened with a multiplex genus-specific primer panel

| Ticks species | Kindia | Mamou | Faranah | Kankan | N'Zerekore | Total |
| --- | --- | --- | --- | --- | --- | --- |
| *Amblyomma variegatum* | 58 | 14 | 22 | 16 | 3 | 113 |
| *Hyalomma truncatum* | 0 | 0 | 2 | 0 | 0 | 2 |
| *Rhipicephalus microplus* | 0 | 0 | 0 | 11 | 0 | 11 |
| *Rhipicephalus geigy* | 1 | 1 | 0 | 9 | 0 | 11 |
| *Rhipicephalus annulatus* | 0 | 0 | 0 | 0 | 1 | 1 |
| *Rhipicephalus (Boophilus) spp.* | 14 | 11 | 0 | 4 | 0 | 29 |
| Total | 73 | 26 | 24 | 40 | 4 | 167 |

In the second stage of the study, we specified the search and screened the entire sample by self-developed qPCR for the detection of the Bunyamwera virus and Ngari virus. All positive samples were sequenced with primers flanking the Bunyamwera virus-specific fragment on the S segment and the Batai virus-specific fragment on the M segment for additional confirmation of the virus species identification (see below for a more detailed description).

### Tick collection

We collected ticks from freshly slaughtered cows or those being prepared for slaughtering in slaughterhouses. Ticks collected from each cow were placed in a separate tube. The sampled ticks were identified to stage and species according to their morphological characteristics (Walker et al. 2003) and pooled into groups of one to five. Pools were formed according to species, sex, and animal host. The ticks were later washed with 70% alcohol and then rinsed twice with 0.15 M NaCl solution. All ticks were stored alive up to the step of the washing in ethanol. Before the washing, we placed tubes with pooled ticks at minus 20°C for 10 minutes for anesthesia. After homogenization, tick suspensions were stored at minus 70° C.

### RNA extraction

Each pool was homogenized with FastPrep-24 (MP Biomedicals, Santa Ana, California, USA) in 1 ml of 0.15 M NaCl solution. Thereafter, DNA and RNA were extracted from 100 μl of the tick suspension using a commercial AmpliSens RIBO-prep kit (Central Research Institute of Epidemiology, Moscow, Russia) following the manufacturer’s instructions. To check the tick species diagnoses based on their morphological characteristics, we sequenced the COI gene fragments for several voucher specimens for each species using the primers jgHCO2198 and ST-COI-F2 (Geller et al., 2013; Makenov et al., 2019). The cDNA was obtained by reverse transcription reactions involving 10 μl of extracted RNA using a REVERTA-L RT kit (AmpliSens, Central Research Institute of Epidemiology, Moscow, Russia), according to the manufacturer’s instructions.

### Screening with a multiplex genus-specific primer panel

We screened 167 tick pools (Table 2) for arboviruses with genus-specific primers according to the approach described by Ayginin et al (2018). The following virus genera were selected for this study: *Orbivirus, Orthonairovirus, Flavivirus, Alfavirus, Phlebovirus*, and *Orthobunyavirus*. The selection of primer sequences for use, preparation of the amplicon libraries, and bioinformatics analysis were the same as those described in the study by Ayginin et al (2018). Sequencing was carried out on the Ion S5 platform using an Ion 520/530 Kit-Chef reagent sample preparation kit and employing Ion 530 chips on the Ion Chef instrument. (Thermo Fisher Scientific, Austin, TX, USA).

### qPCR screening and sequencing

We performed qPCR specific to Bunyamwera virus and Ngari virus with the primers NgvS-F2, NgvS-R2, and probe NgvS-proba2 (see sequences in Table 3). The primers flanked a fragment on the L segment. A total of 710 tick pools were screened with these primers. For each qPCR-positive samples, a fragment of the nucleoprotein gene on the S segment and a fragment of the glycoprotein gene (G2) on the M segment were amplified with the primers listed in Table 3. Subsequent sequencing was conducted using a BigDye Terminator v1.1 Cycle Sequencing Kit (Thermo Fisher Scientific, Austin, TX, USA) on an Applied Biosystems 3500xL Genetic Analyzer (Applied Biosystems, Foster City, CA, USA). The sequences obtained were deposited in the NCBI GenBank database under the following accession numbers: MT748001 – MT748004 (S segment), MT748005 – MT748008 (M segment).

**Table 3.**
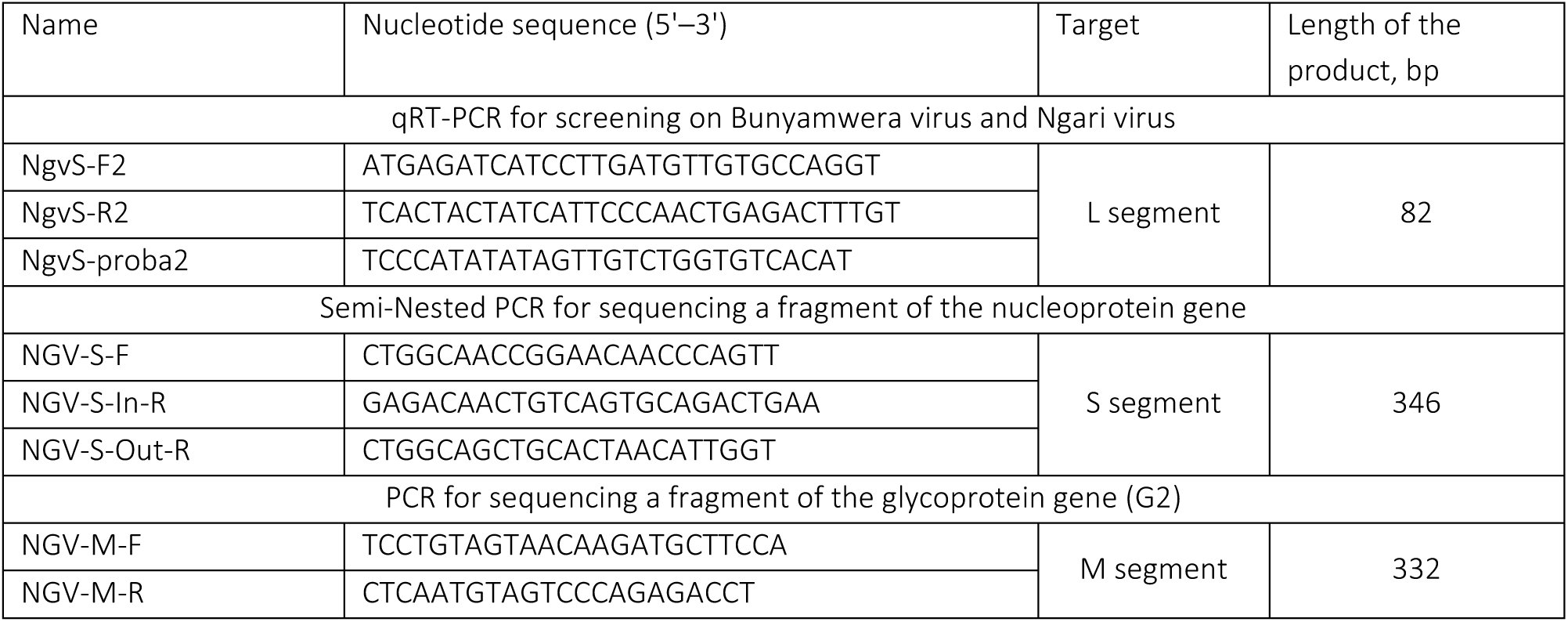
Oligonucleotide sequences of the primers and probes used in PCR amplification and sequencing

### Data analysis

The sites where the ticks were collected were mapped using the free software Quantum GIS. The nucleotide sequences obtained by Sanger sequencing were aligned, compared, and analyzed using ClustalW and BLAST. The Mega X package (Kumar et al., 2018) was used for phylogenetic analyses. Phylogenetic trees were constructed using the maximum-likelihood method with the Tamura 3-parameter model (Tamura, 1992), with bootstrap analysis based on 1000 replicates.

## Results

### Screening with a multiplex genus-specific primer panel

The screening revealed positive results only with primers specific to *Orthobunyaviruses* (L segment). Four pools (N=167) yielded specific amplicons. These pools included adult ticks *Am. variegatum* (five males per pool) collected from the same cow in a village in the Kindia region. Bioinformatic analysis showed that the obtained sequences correspond to the Ngari virus (L segment), with the highest nucleotide sequence identity (∼ 96%) to that of the Ngari virus strain Adrar (GenBank accession KJ716850.1). Based on these findings, we focused our study further on Ngari virus screening.

### qPCR on Ngari virus

During qPCR screening of the whole sample (N=710), we found RNA of Bunyamwera/Ngari virus (L segment) in 12 tick pools that were formed from ticks collected from three different cows in three different villages of the Kindia and Kankan regions (Fig. 1). The average Ct value was 30.5 (SD = 1.2), which indicates a low viral load. Viral RNA was detected in ticks of the following species: *Am. variegatum, Rhipicephalus geigyi*, and *Rh*. (*Boophilus*) spp. Tick species identification was confirmed by sequencing of a fragment of COI-gene (GenBank accession MT107444, MW193715-MW193719). We assume that ixodid ticks are not competent vectors for Bunyamwera virus or Ngari virus and that the PCR-positive ticks had acquired the virus from the blood of infected hosts. Therefore, we did not calculate the prevalence of the virus in ticks but calculated the number of cows from which the Ngari positive ticks were collected (Table 4).

**Table 4.** Ngari virus in ixodid ticks collected from cows in Guinea

| Administrative regions | Number of studied pools | Number of Ngari-positive pools | Number of cows* | Number of Ngari-positive cows** (%–CI 95%) |
| --- | --- | --- | --- | --- |
| Kindia | 199 | 6 | 71 | 1 (1.41 – 0.04-7.60) |
| Boke | 39 | 0 | 13 | 0 (0.00) |
| Mamou | 30 | 0 | 14 | 0 (0.00) |
| Faranah | 182 | 0 | 17 | 0 (0.00) |
| Kankan | 87 | 6 | 32 | 2 (6.25 – 0.77-20.81) |
| N'Zerekore | 173 | 0 | 50 | 0 (0.0) |
| Total | 710 | 12 | 197 | 3 (1.52 – 0.32-4.39) |

\* – the number of corresponding cows from which the studied ticks were collected
\*\* – here “Ngari-positive cows” means cows from which the Ngari positive ticks were collected

To confirm the identification of Ngari virus, we sequenced both the S and M segments of positive pools for each infected cow (Fig. 2). The obtained sequences were aligned with sequences of Bunyamwera virus, Batai virus, and Ngari virus downloaded from the GenBank database. Phylogenetic analyses clearly indicate that the detected viruses are reassortants, with their S segment originating from Bunyamwera virus and their M segment originating from Batai virus (Fig. 2). The sequences are most similar to the sequence of the strain Adrar (KJ716848), which was isolated from goats in Mauritania in 2010 during the Rift Valley fever outbreak (Eiden et al., 2014).

**Fig. 2.**
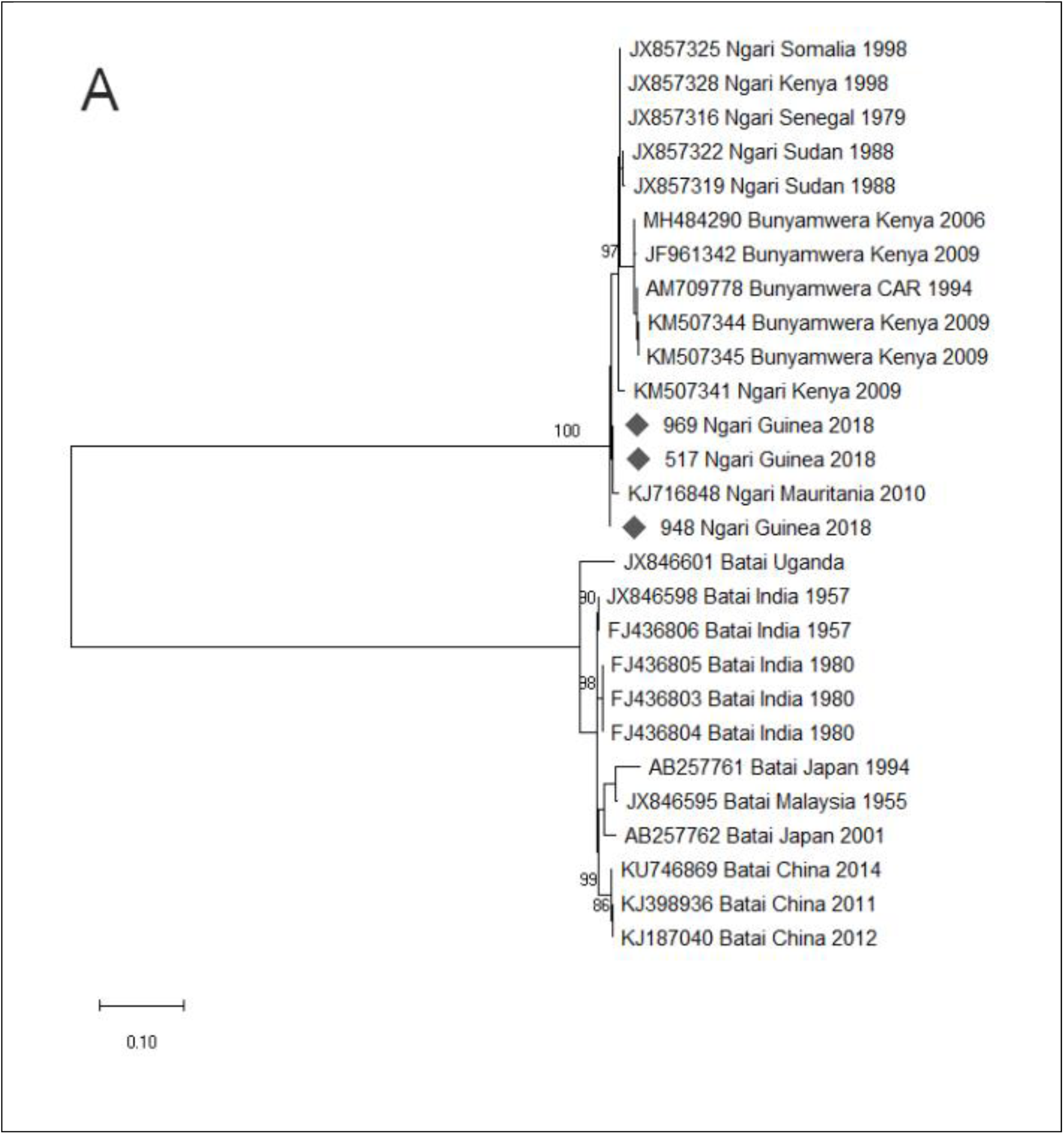
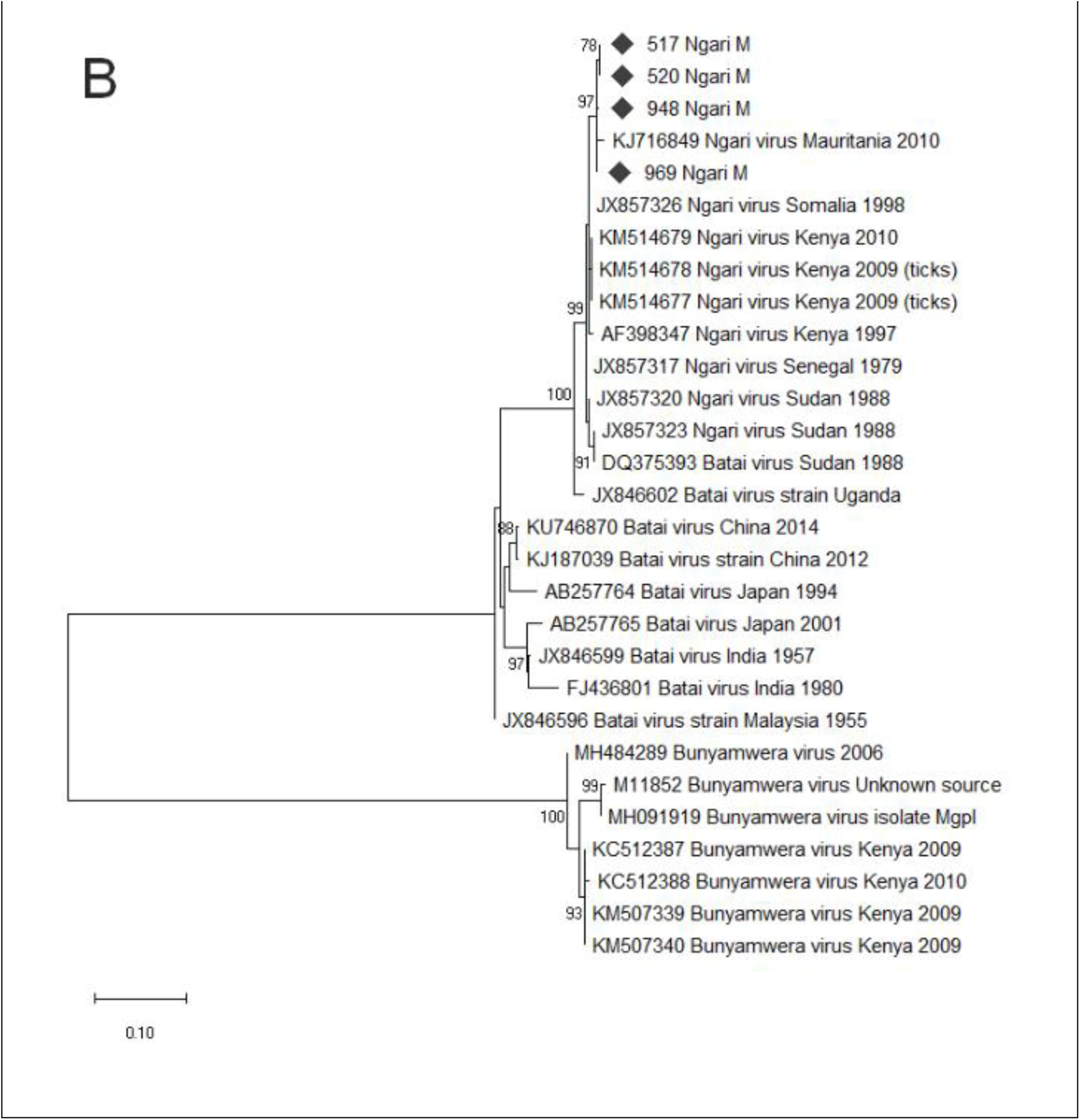
Phylogenetic tree of sequences of Ngari virus-derived A) fragments of the S segment (346 bp) and B) fragments of the M segment (330 bp) of Bunyamwera virus and Batai virus compared with those of isolates obtained from ixodid ticks in Guinea in 2018 (filled diamonds). The trees were constructed by using the maximum-likelihood method (1,000 bootstrap replications) and the Tamura 3-parameter model. A discrete gamma distribution was used to model evolutionary rate differences among the sites. The trees are drawn to scale, with branch lengths corresponding to the number of substitutions per site.

## Discussion

We found Ngari virus in engorged ixodid ticks collected from livestock. Our findings are not the first detection of a mosquito-borne virus in ticks. Previously, cases of Ngari virus isolation from tick species *Amblyomma gemma* and *Rhipicephalus pulchellus* were reported in Kenya (Odhiambo et al., 2016). Other mosquito-borne viruses have also been detected in engorged ticks, including Semliki Forest virus (*Alphavirus*, Togoviridae) in *Amblyomma lepidum, Am. gemma*, and *Rh. pulchellus* (Lwande et al., 2013) and West Nile virus in *Hyalomma marginatum* (Kolodziejek et al., 2014; L’vov et al., 2002), *A. gemma, Rh. pulchellus* (Lwande et al., 2013), and *Ixodes* spp. (Moskvitina et al., 2008; Yakimenko et al., 1991). An exception is the yellow fever virus, which was found in the eggs and larvae of the tick *Am. variegatum* (Germain et al., 1979), for which the transmission of the virus by this tick species was demonstrated in an experiment (Cornet et al., 1982). Bunyamwera virus, which is closely related to Ngari virus, was also isolated from ixodid ticks, including *Am. lepidum, Am. gemma, Rh. pulchellus, Hyalomma truncatum* (Lwande et al., 2013). However, there is no evidence that ixodid ticks are competent vectors of the Ngari virus. In all cases of mosquito-borne virus detection in ticks in Africa listed above, the viruses were isolated from engorged ticks collected from cows or wildlife ungulates. Most likely, ticks obtain such viruses from the blood of an infected host. Studies of the volume of blood ingested by ticks showed that females of *Amblyomma americanum* and *Amblyomma maculatum* obtained up to 1.1 and 2.3 ml of blood, respectively (Koch et al., 1974; Sauer and Hair, 1972). Therefore, engorged ticks might contain not only pathogens for which they are competent vectors but also other pathogens that circulate in the blood of the host at the time of tick feeding.

Our study revealed a low load of viral RNA in ticks, which agrees with the assumption that ticks obtained the virus from infected blood. We found a low prevalence of the virus in cows in two different regions of Guinea. However, here, it is important to note that our estimation of the virus prevalence is biased. The first source of bias involves the study of ticks that are non-competent vectors for the Ngari virus. The efficiency of transmission of Ngari virus from a host with a systemic infection to an adult tick is unknown; therefore, the obtained prevalence of the virus in cows may be underestimated. The second limitation of our results involves the non-representative sample of cows: this study was aimed at the molecular screening of ticks with vector-borne pathogens. We did not consider the number or distribution of cows when designing the sample. However, despite the biased assessment of the prevalence of Ngari virus in cows, our findings allow us to ascertain the presence of the virus in at least two regions of Guinea. For correct estimation of the virus prevalence, further studies of mosquitoes and livestock are needed. In consideration of the detection of other Orthobunyaviruses in ticks and livestock in Guinea, further investigations involving genus-specific primers are needed.

## Data Availability

The authors confirm that the data supporting the findings of this study are available within the article.

## Declarations

### Ethics approval

